# Factors influencing participation in sports, exercise, and physical activity in adolescents with idiopathic scoliosis: a protocol for a qualitative secondary data analysis

**DOI:** 10.1101/2023.07.21.23292992

**Authors:** S. Tucker, A. Soundy, S. Alamrani, A. Gardner, A. Rushton, D. Falla, N.R. Heneghan

**Affiliations:** University of Birmingham; University of Tabuk; The Royal Orthopaedic Hospital NHS Foundation Trust; Western University

## Abstract

**Introduction:** Adolescent idiopathic scoliosis (AIS) is one of the most common paediatric spinal complaints (2-3% of children < 16 years). Regular physical activity is recommended and has been associated with significant improvements in quality of life, reduced pain, and improved function in AIS. However, participation rates remain low amongst individuals with AIS with limited research examining why. This qualitative study aims to identify factors influencing participation in sports, exercise, and physical activities in AIS.

**Methods and analysis:** A qualitative interpretive hermeneutic phenomenology study will be conducted. This study will use a subtle-realist view to enable a focus on the most common experiences of individuals with AIS considering factors influencing participation in exercise, sports, and physical activity. This will be a secondary data analysis (SDA) of a single centre qualitative study completed at a tertiary scoliosis centre during 2022. Participant data drawn from semi structured interviews of individuals <18 years old with a diagnosis of AIS will be included in a six-phase thematic analysis. Rigor will be enhanced through a qualitative checklist, reflexivity, researchers with expertise in the phenomena of interest, and additional researchers from the parent study to critique. Patient and public involvement has been utilised since conceptualisation to improve transparency of reporting.

**Ethics and dissemination:** Full ethical approval was given for this SDA and the parent study by the Health Research Authority (IRAS 289888) and Health and Care Research Wales approval (REC reference: 21/WM/0076). Dissemination will be via peer reviewed publication and conference presentation with results being used to inform future research projects.

**Registration details:** No prior registration has been used for this protocol due to the empirical nature of this secondary data analysis.

**Strengths and Limitations:**

- This project will be conducted with a multidisciplinary research team with expertise in qualitative research, spinal conditions and AIS, and patient and public involvement.
- This study has ethical strengths in its efficiency of data capture from a single parent study. This is significantly cheaper with standardised procedures, less requirement on children suffering from AIS, and relieves the burden of further participant recruitment.
- The nature of SDA requires researcher reflexivity and involvement of the primary researcher from parent study to ensure that there is no loss of contextual information or a lack of immersion in the data.
- The sample will be limited to a single centre study with sampling limited to specialist scoliosis clinics giving opportunity for inferential generaslisation, but lacking opportunity for broad basis generalisations or subgroup analysis.
- The primary aim of parent study was to assess the content validity of the SRS-22r rather than assessment of factors influencing exercise. However, the interview topic guide was reviewed by the secondary data analyst as well as the wider research team to ensure the capture of appropriate and relevant data for this study.

## Introduction

AIS is one of the most common paediatric spinal conditions present in approximately 2-3% of those under age 16 years (1). According to the Scoliosis Research Society, idiopathic scoliosis is a lateral curvature of the spine > 10 degrees Cobb angle combined with rotation and unknown aetiology (2). Of those with AIS some will develop spinal pain, although the exact relationship remains unclear and does not appear to be linked with biomechanics or extent of the Cobb angle (3). Approximately 1 in 5 children with spinal pain experience increasing pain from the age of 6 to 17 years which has a clear association with societal costs (4). Furthermore, children who suffer from prolonged spinal pain are more at risk of developing chronic pain into adulthood (4). AIS presents several key issues including physical deformity, reduced quality of life, and issues surrounding self-perception and self-image (5).

Several management strategies for AIS have been well researched and implemented. Surgery for AIS has been shown to improve both self-image and satisfaction with treatment (6). Furthermore, patients with unoperated scoliosis have reported issues with body image, self-perception and back pain at a 50-year follow-up (7). The literature also suggests that in AIS negative self-perception of body image has a strong correlation with back pain due to the psychological distress (3). Another well researched AIS management strategy is exercise. Although sports, exercise, and physical activity are often used interchangeably they each describe different concepts (8). Sport is described as ‘an organised subset of exercises with a defined goal, it can be spontaneous or organised by specific categories, including but not limited to; age, gender, weight, goal’, etc (9, 10). Although exercise shares several components with physical activity it is not synonymous due to its planned, structured, and repetitive nature (8). Physical activity is a term adopted globally to describe any bodily movement that results in energy expenditure and can include leisure, exercise, or household tasks (8, 11). The International Classification of Functioning, Disability and Health: Children and Youth version (ICF-CY) incorporate a wide range of physical activities into the classification of mobility ranging from basic movements such as changing from lying to sitting to more complex activities such as jumping and swimming (12). Sports, exercise, and physical activity each have a requirement on physical function in order to participate. However, there remains a substantial lack of research regarding factors influencing participation in either sports, exercise, or physical activity in AIS. Therefore, all three components have been searched and explored to determine a meaningful conclusion regarding participation within AIS.

Current UK government exercise guidelines state that children and adolescents over the age of 5 years should participate in 60 minutes of physical activity per day (13), compared with 20 minutes of physical activity per day for those with disabilities (14). There are many benefits to active participation in sports, exercise and physical activity for adolescents with spinal pain (15, 16). Specifically within AIS these benefits include improved quality of life (5, 17). However, the benefits of exercise on Cobb angle continues to be debated (5, 17). Therefore, due to the known benefits of exercise in AIS it is timely to evaluate treatments such as exercise and factors influencing participation (18).

It has been shown that radiographic changes poorly correlate with health-related quality of life (HRQOL) in adults, but treatments such as exercise, that aim to improve pain and promote function are best placed to improve HRQOL (19). Cochrane review evidence identifies no risks associated with participating in exercise in AIS (20). There is some evidence to suggest no difference in restrictions towards sporting activity following spinal fusion compared to nonoperative treatment (21). Whilst the majority of routine practice encourages exercise following spinal surgery for AIS, individuals are usually advised to avoid contact sports post-operatively (22–24). Individuals with AIS who regularly participate in physical activity have improved HRQOL, physical and mental wellbeing, and self-esteem (6, 17, 25, 26). Moreover, adolescent sports participation promotes friendship development and cooperation with others (27). The literature consistently concludes that individuals with scoliosis who regularly participate in exercise have enhancements in HRQOL (28, 29).

Despite the known benefits of exercise for AIS, there remains an association between AIS, back pain, and discontent with body image (30). Furthermore, those with AIS report that their condition limits their participation in sports, exercise and physical activities due to functional impairment and back pain, reducing HRQOL (21, 27). The factors influencing sports participation in AIS patients remains unreported (27). To the best of the authors knowledge only two studies exist within AIS looking at factors influencing physical functioning rather than sports, exercise, and physical activity. One study focused on aspects of physical functioning such as cardiovascular function and immunological function (31). Results identified impaired body structure as a barrier to participation, whilst external support from others acted as a facilitator but did not represent participant voices or experiences (31). A second study identified pre-surgical experiences of individuals with AIS and identified psychosocial support as a key component in living with their scoliosis (32). Due to the lack of understanding regarding factors influencing participation in sports, exercise, and physical activity in AIS this study aims to examine the interview transcripts as part of a secondary data analysis (SDA) for qualitative factors influencing participation and relevant trends across discrete subpopulations.

## Study objectives

1. To identify individual factors influencing participation in sports, exercise and physical activity for participants with AIS.
2. To determine if there are any trends in participation across discrete sub-populations of AIS, such as curve severity, curve morphology or management strategy.

## Methods and analysis

### Research Design

This protocol has been guided and reported according to the Standards for Reporting Qualitative Research (SRQR) guidelines and the Consolidated criteria for Reporting Qualitative research Checklist (COREQ) (33, 34). This study will be a thematic SDA of qualitative data obtained by another member of the research team (SA) as part of a parent study examining the content validity of the SRS-22r (35).

This SDA provides an objective perspective on factors influencing sports, exercise, and physical activity participation in AIS, due to the reduced emotional connection to the data (36). The potential drawbacks of SDA have been mitigated by including members of the parent study research team in the SDA (37). This included the lead researcher for the primary collection (SA), the guarantor of the data (NH), and those advising from a clinical and academic perspective (AG, AR, DF), in addition to new members of the team with expertise in qualitative data analysis (AS), and physiotherapeutic practice (ST).

### Qualitative approach and research paradigm

A subtle-realist world view will be used to help identify a pertinent reality from different qualitative perspectives against a quality criteria (38, 39). Subtle-realism reflects the nature of the research rather than the beliefs of the healthcare researcher (38, 39). Although some argue that subtle-realism has no true ontological basis and lacking certainty (39). Subtle-realism is increasingly embraced as valuable in healthcare research due to its involvement of subjective perceptions whilst not precluding independent phenomena of objects, relationships and experiences (39–41).

Interpretive hermeneutic phenomenology understands that individual experiences are unique, rich and complex (42). Furthermore, using a quality criteria involving validity and relevance the methodology allows the researcher to use qualitative data to denote a pertinent reality (43). The hermeneutic cycle allows an interplay of interpretation and tradition beyond subjective vagueness or objective interpretation towards an understanding of the context of individual experience (42).

### Researcher characteristics and reflexivity

The primary data collector (SA) is a physiotherapist who has developed a reflexive relationship with the data along with the physiotherapists, academics, and surgeons involved in the project, and their perspective influence on data interpretation (43). By recognising and acknowledging foreknowledge from clinical experience, both the primary and secondary data analysists (SA, ST) are able to relinquish attachments to their current knowledge to develop reflexivity (42, 44). Distance between the lead secondary data analyst (ST) and the participants was maintained due to the nature of a SDA, further improving reflexivity (43). Both researchers are female physiotherapists completing doctorate studies, with a special interest in scoliosis and exercise. Participants had no knowledge of the researcher prior to the interview to help further reduce bias (45). Furthermore, advanced preparation of both the primary and secondary data analyst allows in depth interviewing due to a comprehensive understanding of the context of a respondent’s community, generation, work, and personal family life (37). Furthermore, the SDA will be completed closely following primary data collection helping to ensure accurate interpretation due to the risk of interpreting ability waning over time (37, 46).

### Patient and public involvement (PPI)

PPI has been involved from the conceptualisation stage and will continue to be until dissemination. This study has involved those who are not participants in research decisions including clinicians in the field and academics regarding research methodology. There has been a PPI representative (ER) involved in giving feedback throughout the parent study on the study protocol, participant information sheet, interview topic guide, consent form, and has reviewed original study results. The guidance for reporting involvement of patients and the public (GRIPP2) checklist will be used to further promote quality, transparency, and consistency of PPI reporting in this study, Supplementary File 1 (47).

### Context

The primary data collection took place through interviews virtually over Zoom from January to April 2022. This was done in accordance with national guidance on COVID-19 precautions at the time of study conception (48, 49). Prior to primary data collection, interview questionnaires were co-designed and co-constructed with clinicians working in the field, academics, and the secondary data analyst to ensure relevance of the work being completed (50). One pilot interview was conducted with an individual who has scoliosis age 21; this allowed the primary researcher to balance both researcher relationship and rigor, reformulate and refine questions, and ensure focus as required in healthcare (51–54). No substantial changes to the topic guide were made to the topic guide, Supplementary File 2.

### Sampling strategy

All participants were recruited using purposive sampling from a single tertiary scoliosis centre in the UK during the parent study by a specialist surgeon and member of the research team (AG). Purposive sampling achieved theoretical representativeness whereby the sampling and generalisability of the parent study is translated to the SDA due to similarities in time, place, and participants (55). Although qualitative data is not empirically generalisable, purposive sampling ensures results are theoretically generalisable, a complex process to understand concepts, phenomena, and propositions relevant to other similar settings and individuals (44, 56, 57). To reduce selection bias recruitment occurred independently of the primary researcher or secondary data analyst prior to the interview (45). The sample size (n=11) was deemed sufficiently large and varied for analysis with size being determined by participant contribution (58). Furthermore, sample size was determined by data saturation, which was achieved by interview seven, remaining interviews identified that no further interviews were required. Data saturation was determined during the parent study using a saturation table (35). During the parent study naturalistic generalisation was achieved, this understands that results remain generalisable whilst maintaining individual subjective meaning (59).

### Eligibility Criteria

The population of interest for this study was adolescents with AIS aged 11-18 years. All individuals have scoliosis with a Cobb angle greater than 10 degrees and no maximum curve size has been applied as an exclusion criteria to participants (60). No other exclusion criteria were applied with regards to gender, language, ethnicity, or socioeconomic status.

### Ethical issues pertaining to human subjects

One of the chief concerns with SDA are the confidentiality agreements. However, ethical approval was gained to ensure the SDA does not exploit participants original consent (36, 37, 61). Analysed data will be anonymised prior to the SDA conforming to legal and ethical guidelines regarding protection of participant identity without losing the value in qualitative information (37).

### Data collection

Demographic data was recorded for each participant including age, gender, ethnicity, curve location and severity, pain intensity, SRS-22r scores, management of AIS, and physical activity levels. The lenke classification was used to describe participants spinal curvature, allowing comparative analysis between participants and treatments (62).

Data were collected using semi-structured interviews with the first researcher over Zoom. Interviews consisted of two parts; firstly ethics statement, background of the SRS-22r, introductory questions, transition questions, and concept elicitation. The second part of the interview consists of cognitive debriefing of the SRS-22r, followed by a conclusion. See Supplementary File 2 for the topic guide. The first section includes questions such as ‘Do you like exercising?’ and prompt questions such as ‘What is it that you like / dislike about exercising?’. This was done within the parent study to identify key concepts regarding the influence of AIS on quality of life (35). The second part of the interview was a cognitive debriefing to assess the content validity of the SRS-22r (35). The second part included questions such as ‘what is your current level of activity?’ providing valuable information for the purposes of our secondary data analysis exploring factors influencing sports, exercise, and physical activity participation in AIS (35).

Interviews were standardised with the first researcher and all eligible participants enrolled. Participants were allowed to choose to complete the interview with a parent, caregiver or guardian. Following interview completion the first researcher (SA) completed analysis for the parent study and the second researcher (ST) will complete the SDA. Interviews were audio recorded and externally transcribed by official transcription services. The secondary data analyst (ST) will complete thematic SDA using clean pre-transcribed interview transcripts and anonymised participant demographic data in the context of subtle realism. As guided by the National Institute for Health and Care Research (63), data will be rigorously handled and stored. All data and information is protected and stored by the research electronic data capture (REDcap) software (64).

### Data collection instruments and technologies

The topic guide (Supplementary File 2) allowed for purposeful exploration of the phenomena of interest whilst simultaneously allowing comprehensive and systematic exploration of participant responses (65). The topic guide was created by the primary data collector (SA) alongside academics with expertise in the field (NH, AR, DF, AG, AS) and the secondary data analyst (ST) to ensure that interview data was relevant and targeted to both the parent study and the SDA.

As per recommendations semi-structured interview questions were designed with open-ended, clear, and neutral language (51). The interview started with simple contextual questions before moving into more in-depth questions to understand participant experience and their perception of the world (51). Questions on the main theme and follow-up questions are used throughout exploring participants thoughts, feelings, and beliefs (51, 54).

The SRS-22r was chosen as a data collection tool to comprise the second half of the interview (66). This outcome measure was chosen due to its internal consistency, reliability and reproducibility (67). Furthermore, the SRS-22r is one of the most commonly used and accepted patient reported outcome measures and has been shown to provide information on HRQOL that is not obtained from other outcome measures such as the Child Health Questionnaire (CHQ-CF87) (66–68). Although the SRS-22r scores do not correlate well with Cobb angle and trunk deformity measurements, it has been shown to be more sensitive towards HRQOL than other outcome measures such as the CHQ-CF87 (67, 68). Further work remains ongoing into content validity testing and a wider application of the SRS-22r (35). Through calculating SRS-22r scores and interviewing participants to obtain verbal accounts that acknowledge biopsychosocial factors we can better understand participation in physical activity, sports, and exercise in AIS.

### Data items and units of study

The full interview comprised semi-structured questions surrounding factors influencing participation in sports, exercise, and physical activity. Following this the second half of the interview is comprised of completion and content validity testing of the SRS-22r for the purposes of the parent study, however qualitative responses were incorporated into the SDA (Supplementary File 2) (66).

### Data outcomes, processing, and prioritisation

Following data collection, transcripts will be analysed for trends in qualitative factors surrounding participation in sports, exercise and physical activity. Throughout the process information on participant demographics and attrition rate will be recorded to allow for internal and external validity, reduce bias, and improve generalisability (69). Due to the nature of patient confidentiality in the National Health Service and individual qualitative interviews we are able to expose unexpected spillover (70, 71). This study was completed in a single setting and therefore it is important to be aware that lack of replication in other settings may influence robustness (71).

The secondary data analyst (ST) will ensure audit trails, critical reflective comparison, ethical practice and rigor is maintained and therefore making it an appropriate form of evaluating factors influencing participation (72).

### Data analysis and synthesis

A six stage Braun and Clarke reflexive thematic analysis (73) will be used to establish trustworthiness (73, 74). This is the same six stage process that was used during thematic analysis of the parent study (35). Firstly, phase one involves data transcriptions which will be thematically analysed for familiarity (73). Phase two involves coding through labelling participant quotes for similarities and differences (73–75). During phase three, data will be searched for themes and grouped together under corresponding codes with all quotations under that code being listed for analysis and review (73–75). Themes identified will be searched for during phase three and then reviewed during phase four (73, 74). This allows for searching across all quotations by certain words or phrases as well as visualising the data on a certain theme (75). During phase four, data will be thematically synthesised using direct quotations to produce descriptive themes (76). Any subthemes identified in relation to demographic data will also be recoded for analysis and synthesis. Phase five involves naming the themes with both themes and narratives will be described in depth to reflect the broad range of experiences discussed in the data and promote transferability (51, 73, 74). This process will involve an in-depth description of multiple participant perspectives and narratives with a traditional integration of quotes into the results text (51). Finally phase six of data synthesis is producing the report, data analysis and synthesis is complete themes, subthemes and participant quotes will be represented in a table to allow for easy visualisation and interpretation of results (73, 74).

### Techniques to enhance trustworthiness, reduce risk of bias, and increase confidence in the evidence

To ensure credibility, other members of the research team will be involved throughout the process giving congruence between respondents’ views and the researchers representation (74). This study will involve peer debriefing throughout to provide an external check on the research process and promote credibility (74). Furthermore, data analysis, systematising and audit process will be documented throughout to help increase rigor (36, 74).

Numerous techniques have been used to help enhance trustworthiness. These include identifying how the nature of secondary data analysis may influence results, increasing rigor and identifying any limitations in qualitative SDA (36). This study endeavours to increase rigor and trustworthiness by involving research team members from the primary data collection with specific expertise (SA, NH, AG, AR, DF) as well as new members with expertise in spinal surgery and research methodology (AS) including the secondary data analyst (ST) with new perspectives uninfluenced by the primary analysis (36). Those completing the SDA will complete it with uncoded transcripts with audit trails and peer debriefing throughout (36). Further identification of limitations in qualitative SDA for example changes in time, societal norms, or context will be identified (36). Beneficially, SDA reduces the researcher’s emotional investment in data and thereby helps objectify results (72). The SDA has been planned from early on in the primary study prior to data collection to ensure that goals and purpose of the secondary data analysis are a good fit with the primary study increasing confidence in the evidence (36).

Quality will be promoted throughout this study using researcher triangulation, participant validation, peer review, negative case analysis, and using multiple data coders with standardised methods (74, 77). This study follows a previous systematic review (78) and has a clear research question with continuity between the aims and methodology (76). Furthermore, the methodology promotes rigor in sampling, data collection, exploration of deviant cases and researcher reflexivity (76). Audit processes and documentation will be used throughout to encourage dependability (74). Due to ethical considerations, resource, and epistemological challenges member checking is not possible in order to ensure participant protection (79). There are no conflicts of interest identified in this protocol and transparency will be promoted throughout (76, 80). This protocol and methodology has been written and guided by both the COREQ and SRQR checklist, both shown to help improve transparency and ensure explicit, comprehensive reporting (33, 34). The Joanna Briggs Institute Checklist for Qualitative Research (JBICQR) will be used to guide the qualitative research and ensure that results are sensitive and valid (81, 82).

## Discussion

Participation in sports, exercise, and physical activity has been shown to have numerous benefits for individuals suffering with AIS including improved quality of life, improved pain, and improved function (5, 17). However, many individuals with AIS do not participate in sports and exercise and there is limited qualitative research examining why (27). There is limited research suggesting possible reasons why asymptomatic healthy children and adults do not participate in sports, such as lack of motivation, skill and feeling self-conscious (83). Comparatively, desire for social interaction and enjoyment helps facilitate participation (83). However, to the best of the authors knowledge, no work has been undertaken to examine qualitative factors influencing participation in sports, exercise, and physical activity specifically within AIS. Furthermore, to determine if there are any trends dependent on demographics such as curve severity or morphology as defined by the Lenke classification, pain intensity, or management stragegy (62).

Factors such as pain and curve severity have been shown as limitations to sports, exercise, and physical activity participation in AIS and reduced quality of life (21, 27). However, the majority of the literature remains surrounding exercise participation in AIS remains quantitative with no understanding surrounding individual choice to participate in or abstain from exercise (21, 27, 84). Further robust literature is needed to examine these factors and better understand how physiotherapists can engage with indivudals diagnosed with AIS to help the individual engage with sports, exercise, and physical activity and promote the benefits of exercise in an individualised, achievable, and sustainable manner (85–87).

There is a small body of evidence surrounding safety of when it is appropriate for an individual to return to sports, exercise, or physical activity following corrective surgery in AIS (23, 24, 88). However, there remains very little information regarding rates of participation and uptake of sports, exercise, and physical activity (21, 27). Furthermore, to the best of our knowledge there is no evidence exploring specific individual opinions, beliefs, or attitudes influencing participation in sports, exercise, or physical activity in AIS in either post-operative or conservatively managed individuals.

Due to the benefits of sports, exercise, and physical activity participation in AIS and the lack of understanding surrounding factors influencing participation, this secondary data analysis aims to identify key factors influencing participation and if there is a relationship with curve severity, lenke type, pain intensity, or management type.

## Ethics and Dissemination

Full ethical approval was provided for the parent study by the Health Research Authority (IRAS 289888) and Health and Care Research Wales approval (REC reference: 21/WM/0076). Further ethical approval for the SDA of interview transcripts for the purposes of this study was provided by the Health Research Authority and Royal Orthopaedic NHS Foundation Trust ROH21ORTH04. Data collected from studies will be stored in accordance with the UoB data protection policy. Data from included studies and data used for analysis can be accessed when requested from the primary author.

The results from this paper will aim be disseminated through both conference presentations at Physiotherapy UK 2023, EuroSpine 2024, the Physiotherapy Research Society 2024, and targeted journal publications. Possible journals include Spine or PlosOne. Additionally, the results will be used to inform future research and provide training for clinicians undertaking evidence-based practice (89).

## Authors’ contributions

Study conceived by NH, AS, AG, AR, DF, SA, ST, with all authors contributing to shaping the design and methods. ST drafted the manuscript with guidance from AS and NH. All authors contributed to the revising and re-drafting the manuscript. The final version was approved by all authors with agreement to submission to BMJ Open. NH is the guarantor of the work, with ST the corresponding author.

## Supplementary File 1: GRIPP 2 CHECKLIST

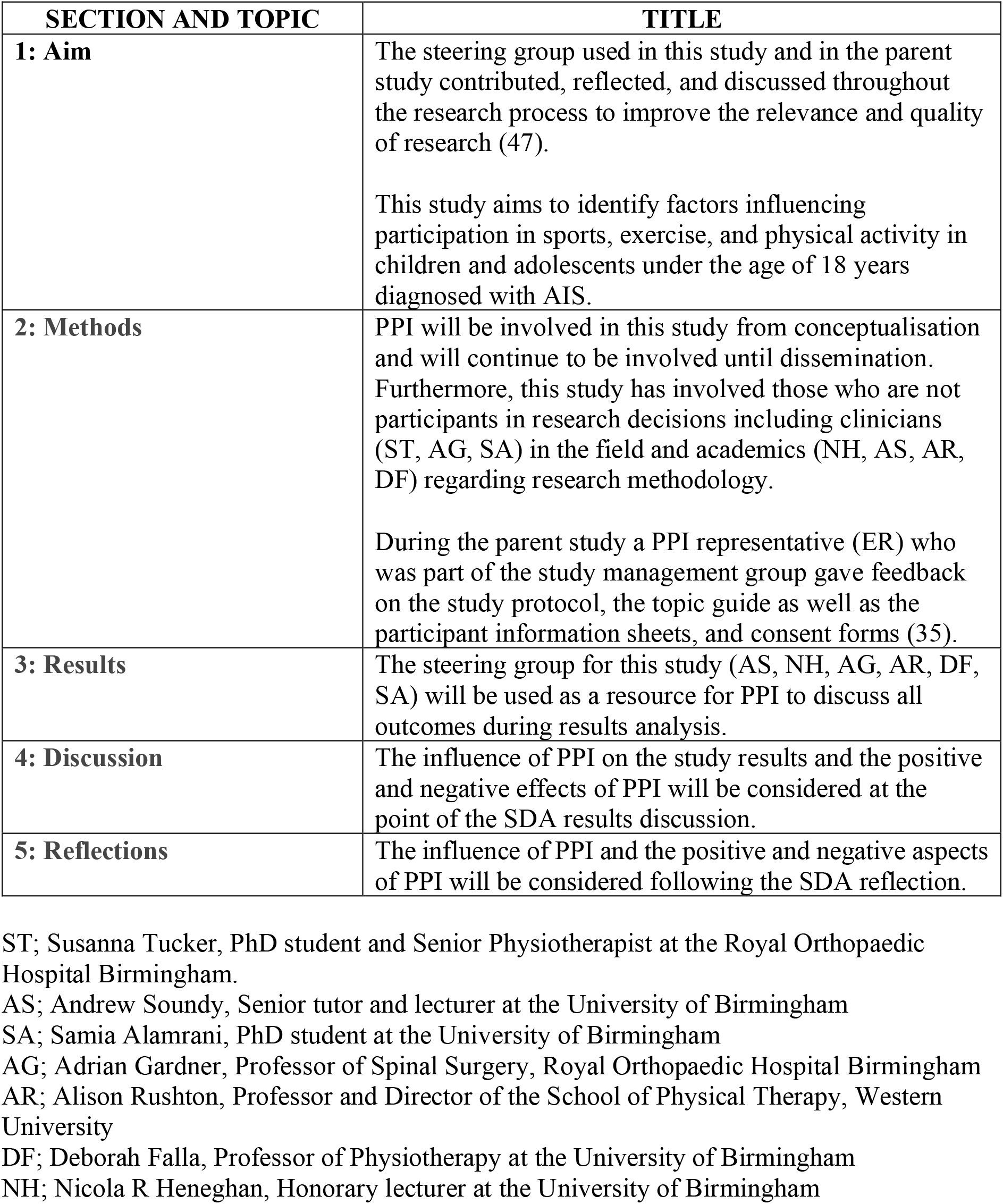

## Supplementary File 2: Topic Guide

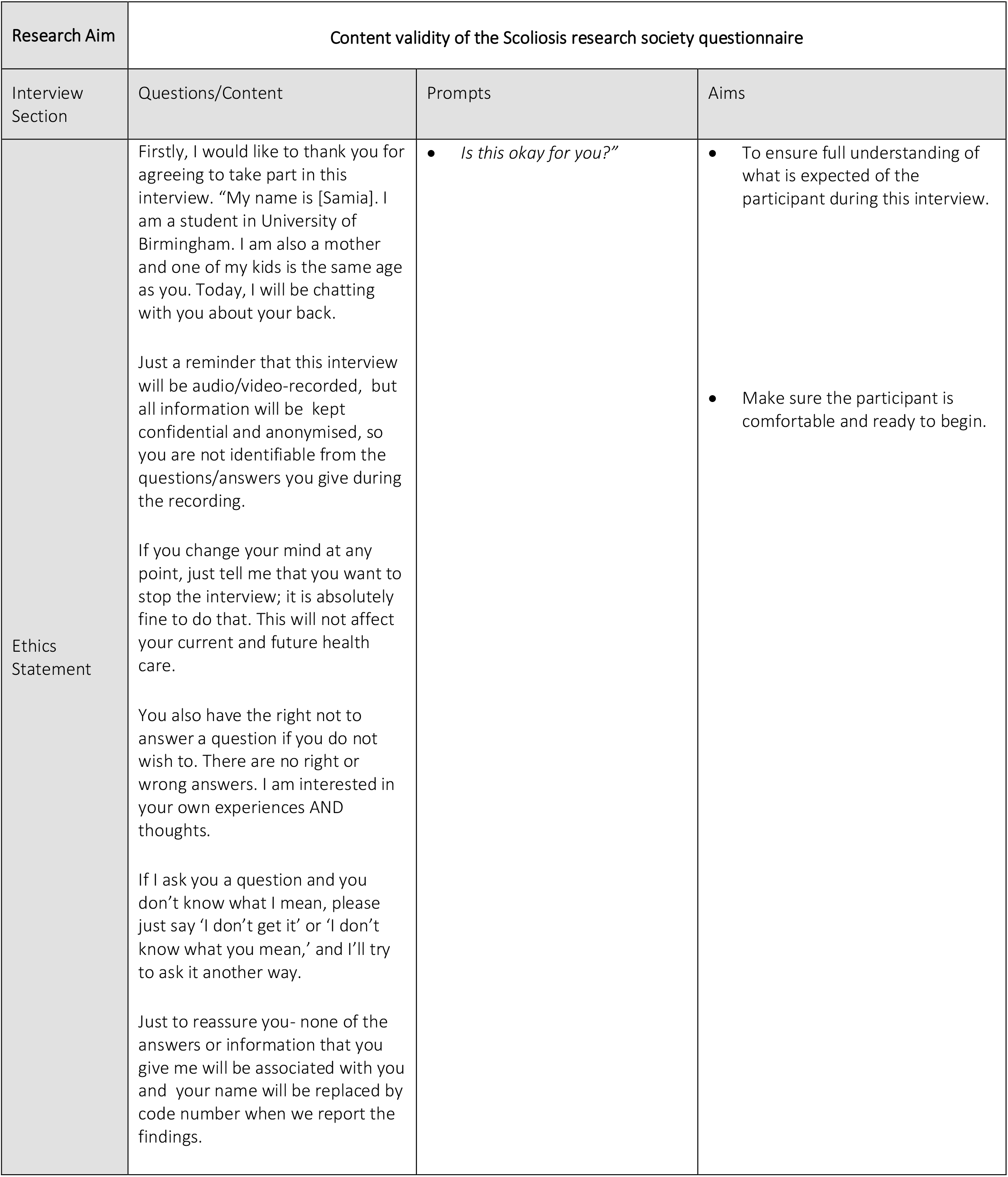
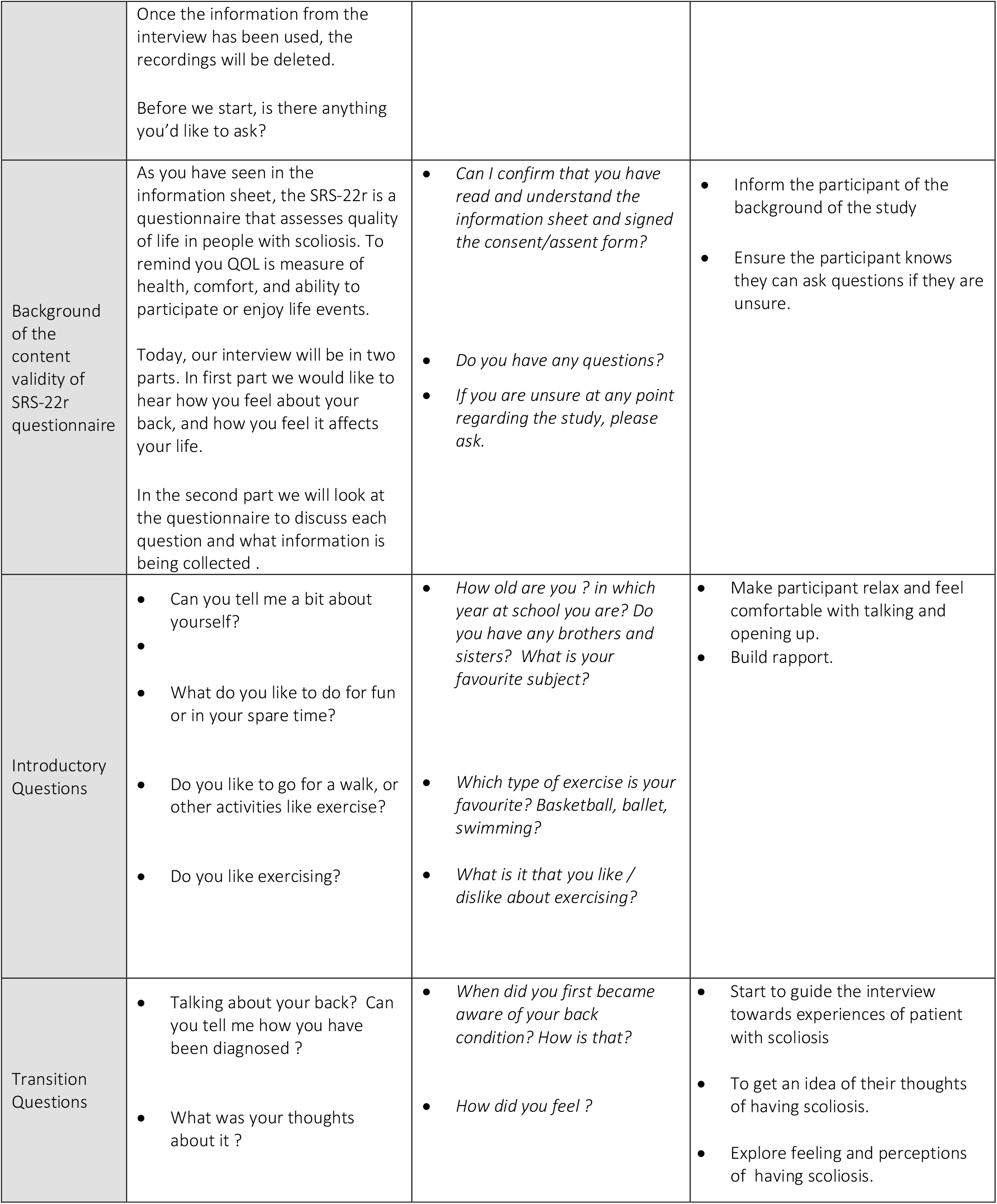
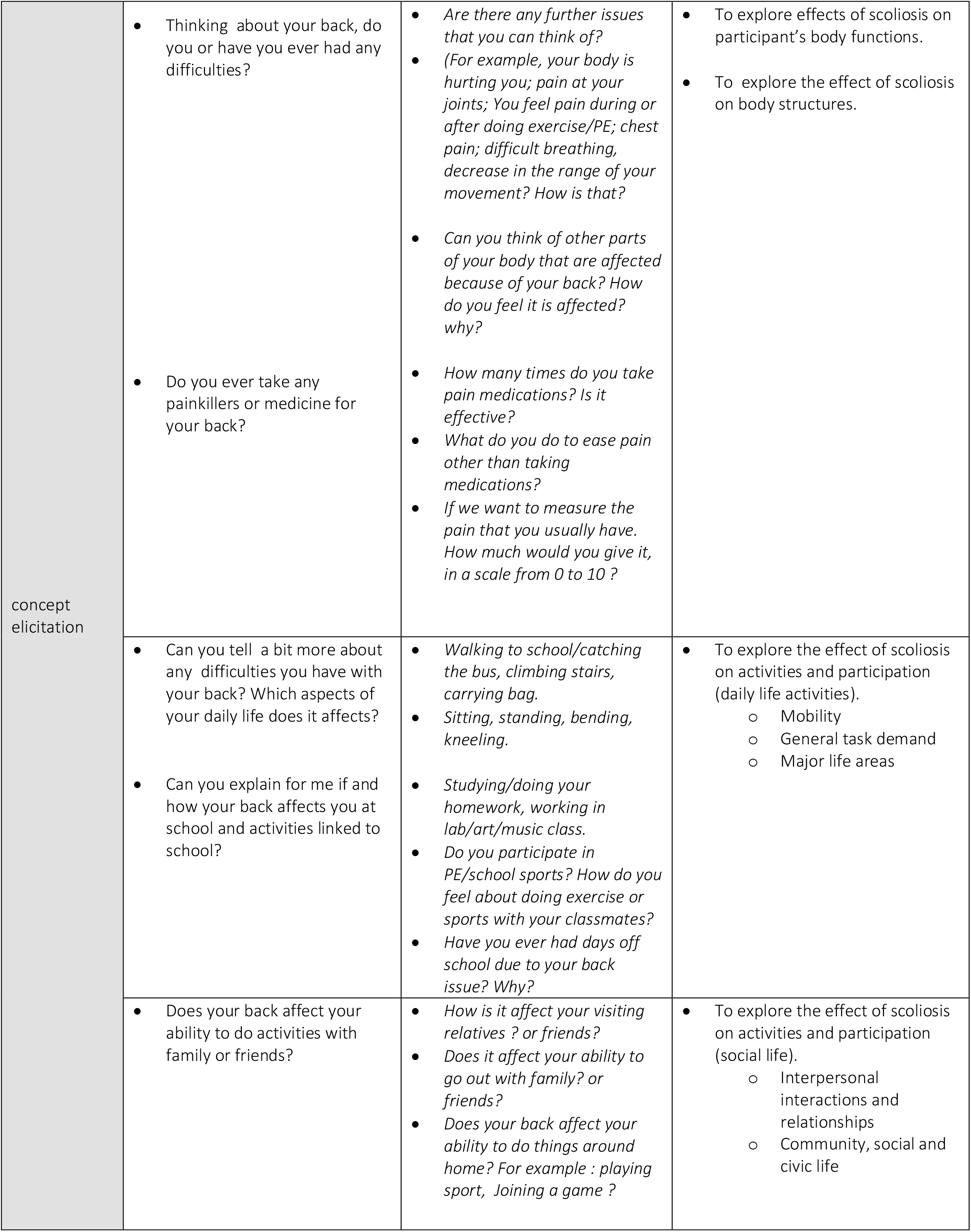
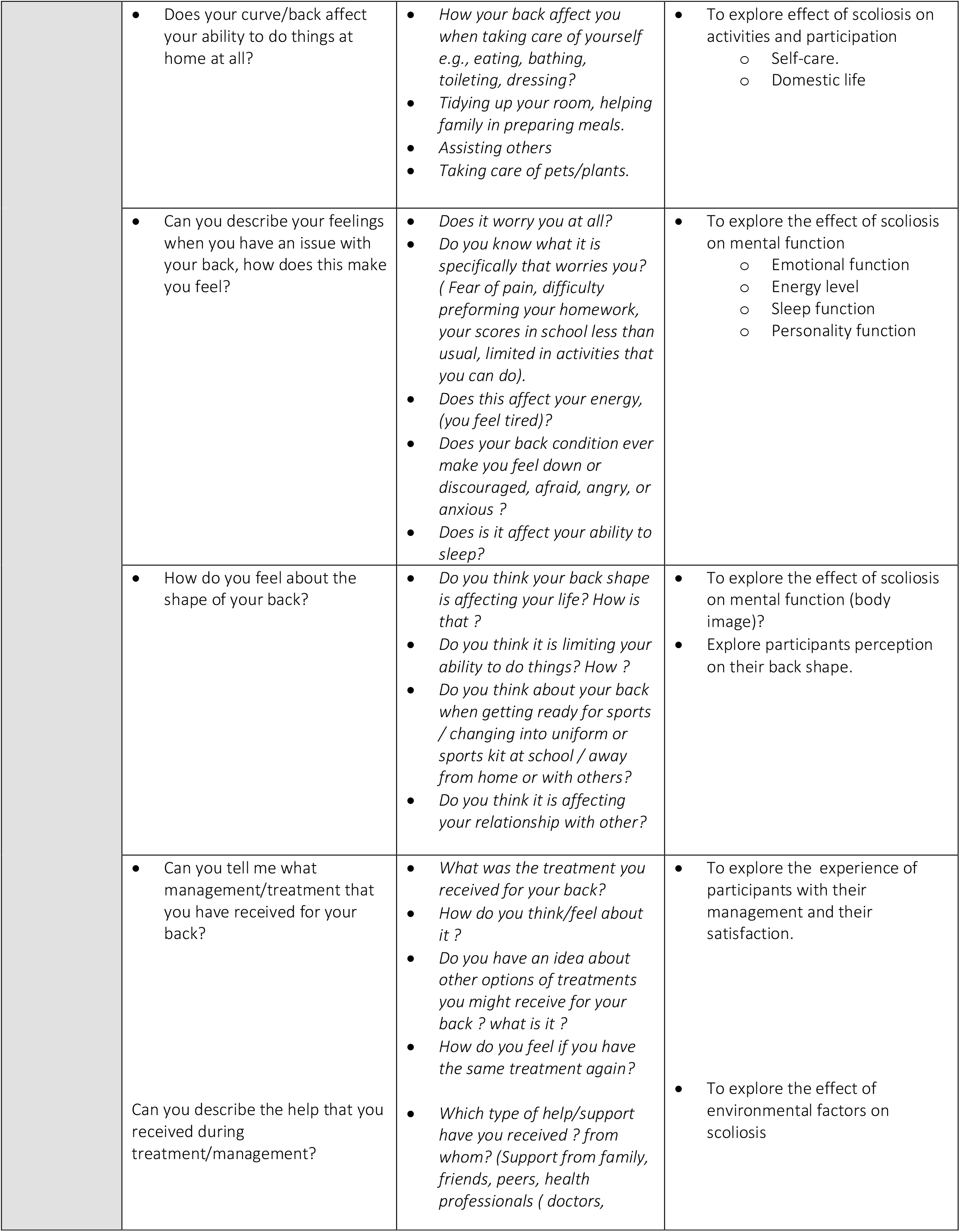
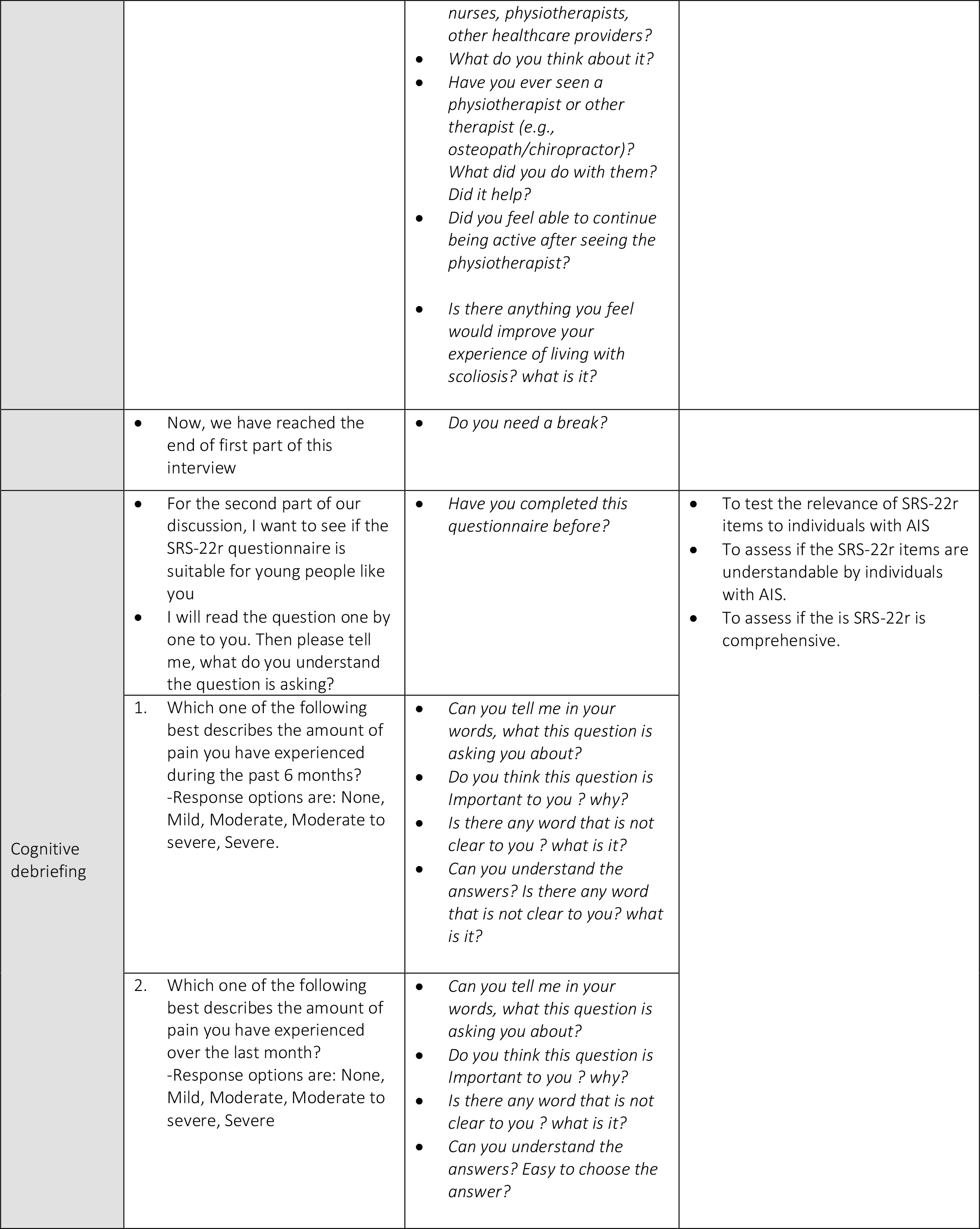
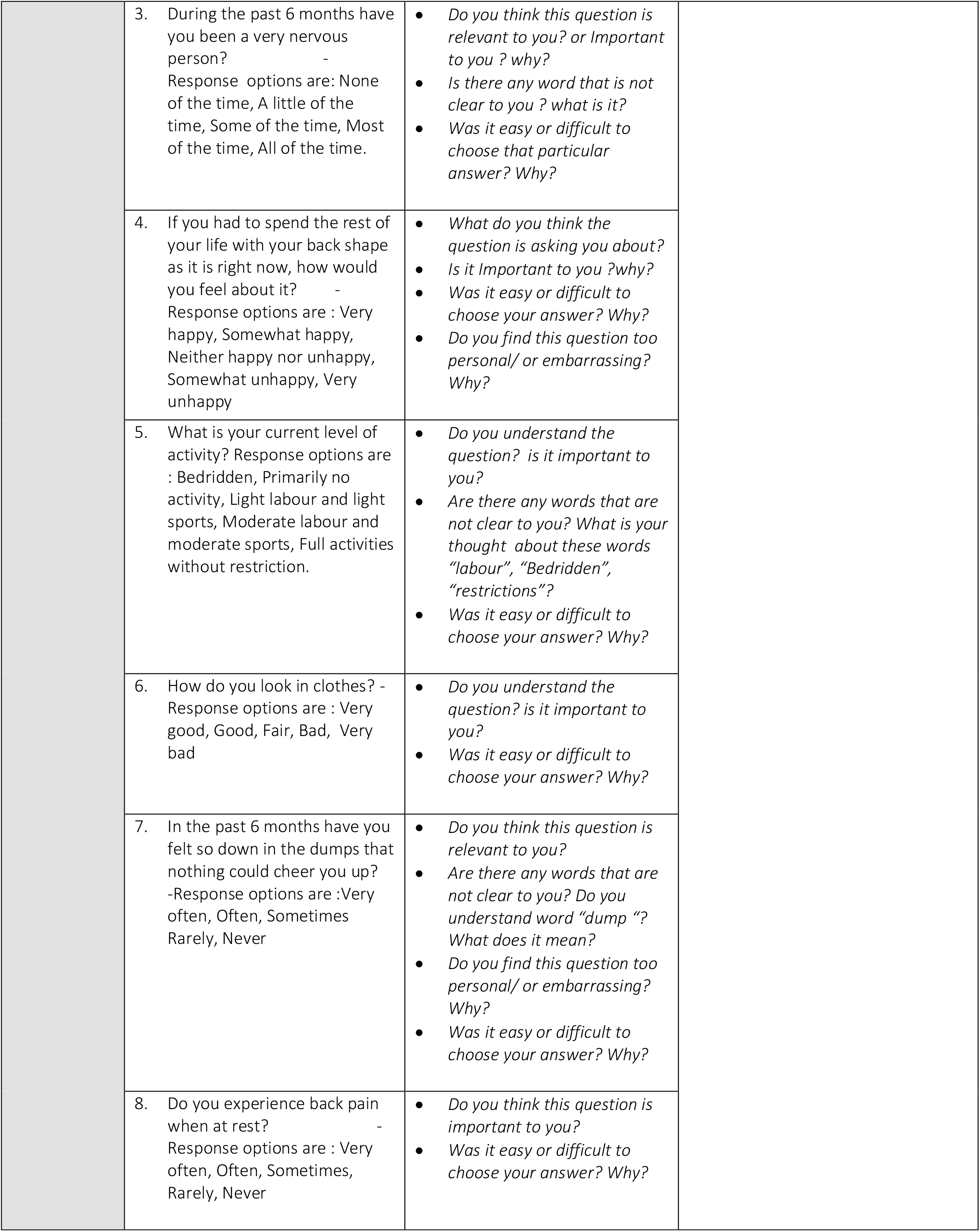
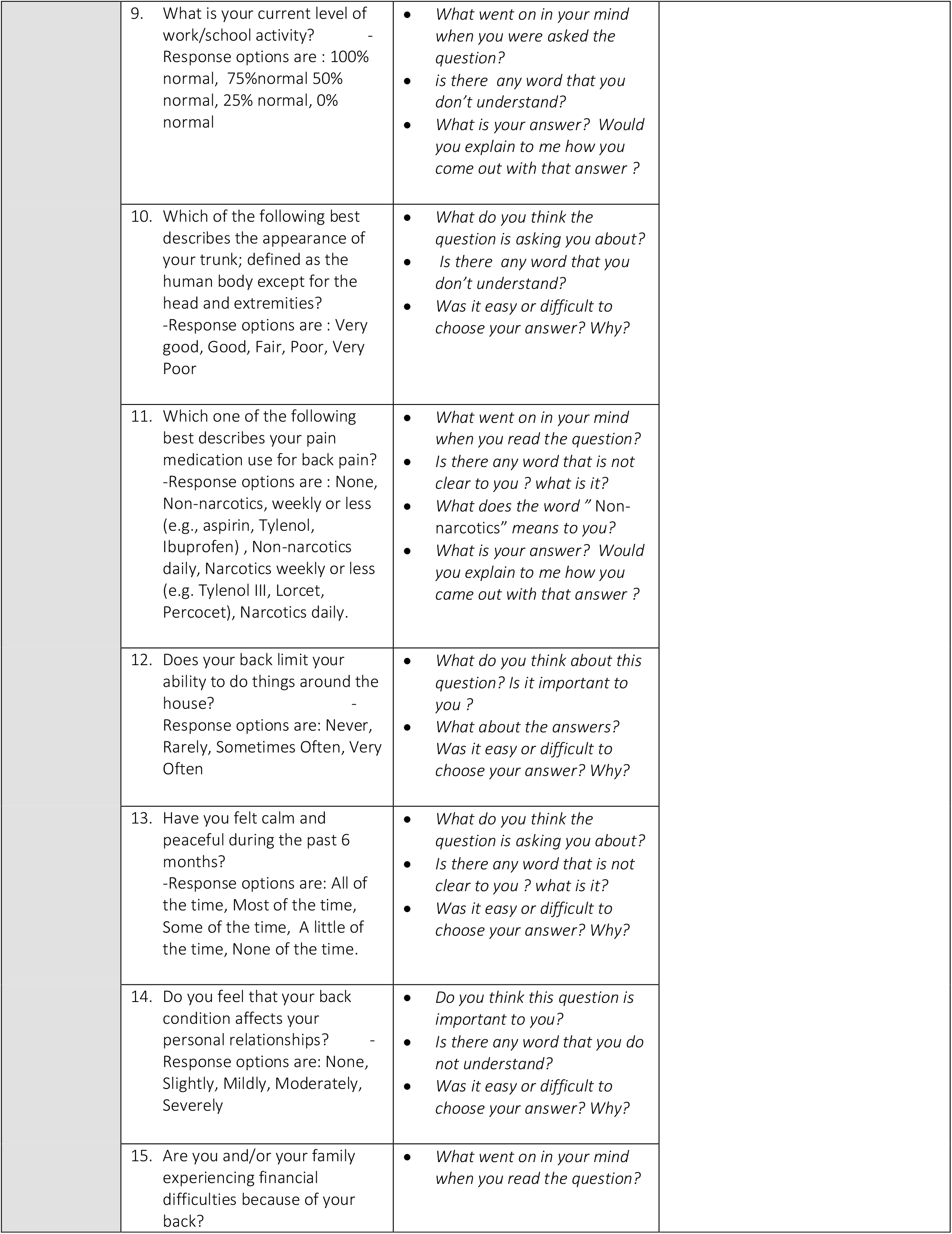
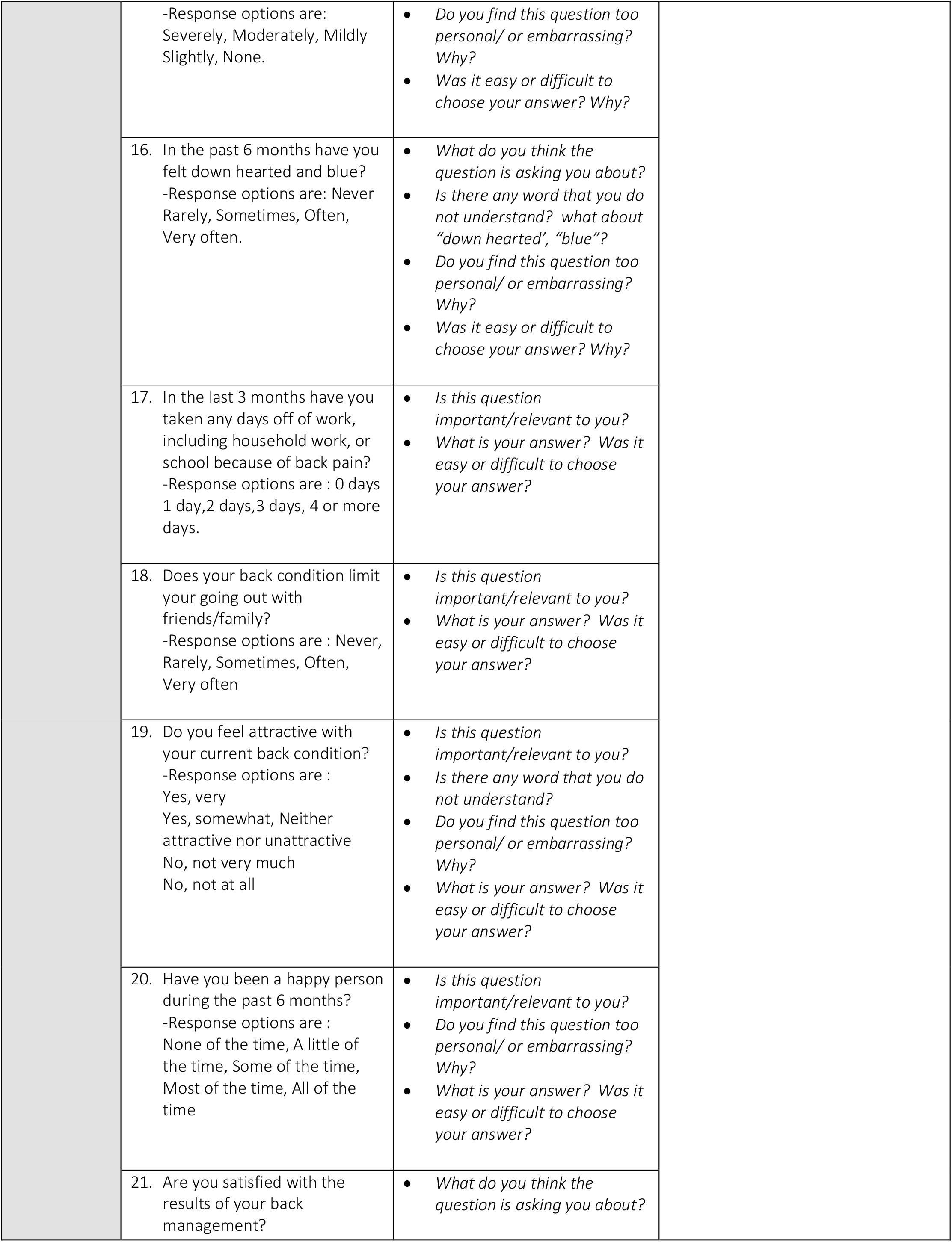
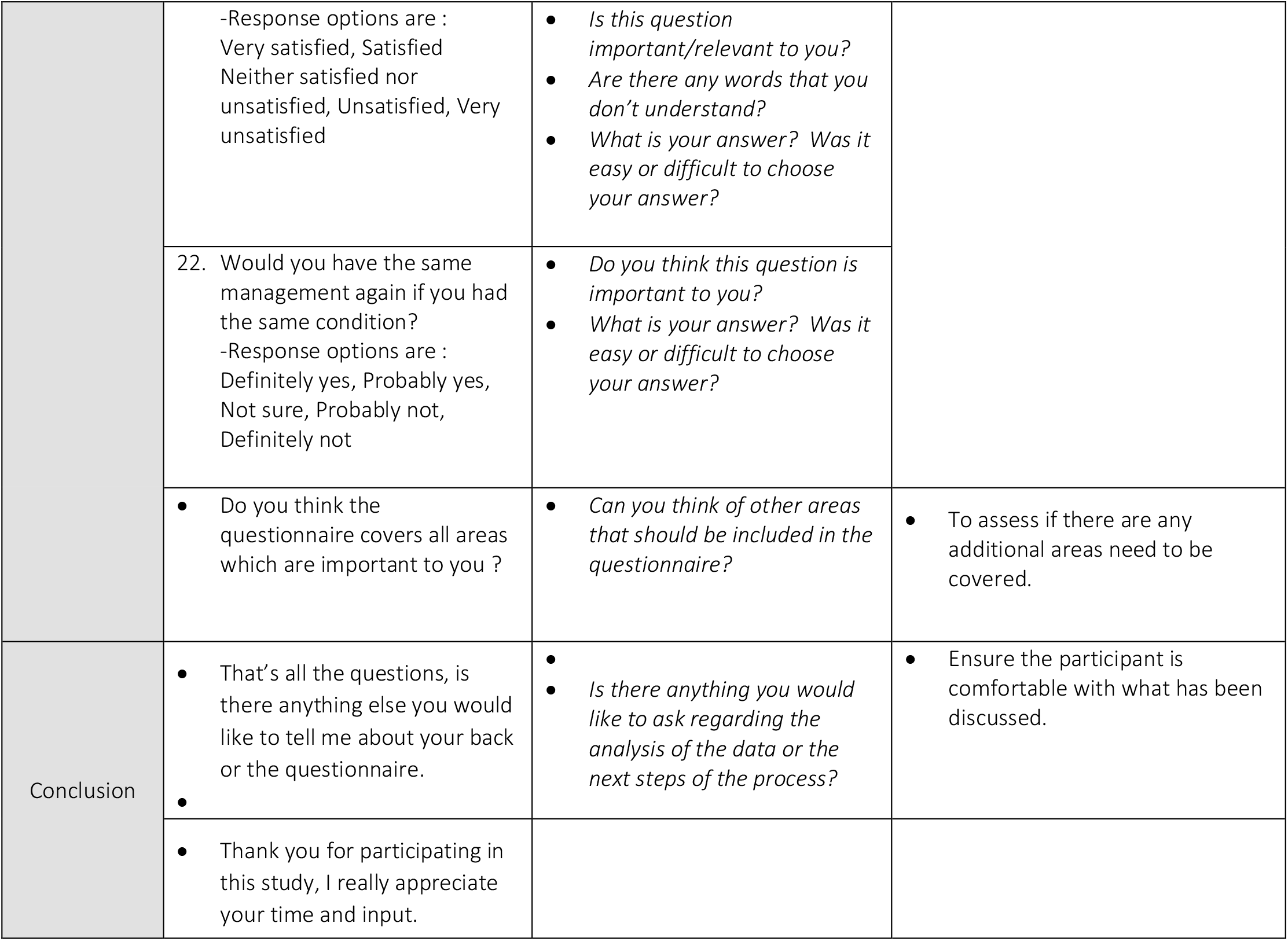

## Data Availability

All data and information is protected and stored by the research electronic data capture (REDcap) software. All data produced in the present study are available upon reasonable request to the authors.

