## Supplemental File 1 and 2 for "Factors influencing participation in sports, exercise, and physical activity in adolescents with idiopathic scoliosis: a protocol for a qualitative secondary data analysis"

**Supplementary File 1:** GRIPP 2 CHECKLIST

| SECTION AND TOPIC | TITLE |
|---|---|
| **1: Aim** | The steering group used in this study and in the parent study contributed, reflected, and discussed throughout the research process to improve the relevance and quality of research (47).<br><br>This study aims to identify factors influencing participation in sports, exercise, and physical activity in children and adolescents under the age of 18 years diagnosed with AIS. |
| **2: Methods** | PPI will be involved in this study from conceptualisation and will continue to be involved until dissemination. Furthermore, this study has involved those who are not participants in research decisions including clinicians (ST, AG, SA) in the field and academics (NH, AS, AR, DF) regarding research methodology.<br><br>During the parent study a PPI representative (ER) who was part of the study management group gave feedback on the study protocol, the topic guide as well as the participant information sheets, and consent forms (35). |
| **3: Results** | The steering group for this study (AS, NH, AG, AR, DF, SA) will be used as a resource for PPI to discuss all outcomes during results analysis. |
| **4: Discussion** | The influence of PPI on the study results and the positive and negative effects of PPI will be considered at the point of the SDA results discussion. |
| **5: Reflections** | The influence of PPI and the positive and negative aspects of PPI will be considered following the SDA reflection. |

ST; Susanna Tucker, PhD student and Senior Physiotherapist at the Royal Orthopaedic Hospital Birmingham.
AS; Andrew Soundy, Senior tutor and lecturer at the University of Birmingham
SA; Samia Alamrani, PhD student at the University of Birmingham
AG; Adrian Gardner, Professor of Spinal Surgery, Royal Orthopaedic Hospital Birmingham
AR; Alison Rushton, Professor and Director of the School of Physical Therapy, Western University
DF; Deborah Falla, Professor of Physiotherapy at the University of Birmingham
NH; Nicola R Heneghan, Honorary lecturer at the University of Birmingham

**Supplementary File 2:** Topic Guide

| Research Aim | Content validity of the Scoliosis research society questionnaire | | |
|---|---|---|---|
| Interview Section | Questions/Content | Prompts | Aims |
| Ethics Statement | Firstly, I would like to thank you for agreeing to take part in this interview. "My name is [Samia]. I am a student in University of Birmingham. I am also a mother and one of my kids is the same age as you. Today, I will be chatting with you about your back.<br><br>Just a reminder that this interview will be audio/video-recorded, but all information will be kept confidential and anonymised, so you are not identifiable from the questions/answers you give during the recording.<br><br>If you change your mind at any point, just tell me that you want to stop the interview; it is absolutely fine to do that. This will not affect your current and future health care.<br><br>You also have the right not to answer a question if you do not wish to. There are no right or wrong answers. I am interested in your own experiences AND thoughts.<br><br>If I ask you a question and you don't know what I mean, please just say 'I don't get it' or 'I don't know what you mean,' and I'll try to ask it another way.<br><br>Just to reassure you- none of the answers or information that you give me will be associated with you and your name will be replaced by code number when we report the findings. | • *Is this okay for you?"* | • To ensure full understanding of what is expected of the participant during this interview.<br><br>• Make sure the participant is comfortable and ready to begin. |

| | | | |
|---|---|---|---|
| | Once the information from the interview has been used, the recordings will be deleted.<br><br>Before we start, is there anything you'd like to ask? | | |
| Background of the content validity of SRS-22r questionnaire | As you have seen in the information sheet, the SRS-22r is a questionnaire that assesses quality of life in people with scoliosis. To remind you QOL is measure of health, comfort, and ability to participate or enjoy life events.<br><br>Today, our interview will be in two parts. In first part we would like to hear how you feel about your back, and how you feel it affects your life.<br><br>In the second part we will look at the questionnaire to discuss each question and what information is being collected . | • *Can I confirm that you have read and understand the information sheet and signed the consent/assent form?*<br><br>• *Do you have any questions?*<br>• *If you are unsure at any point regarding the study, please ask.* | • Inform the participant of the background of the study<br><br>• Ensure the participant knows they can ask questions if they are unsure. |
| Introductory Questions | • Can you tell me a bit about yourself?<br>•<br>• What do you like to do for fun or in your spare time?<br><br>• Do you like to go for a walk, or other activities like exercise?<br><br>• Do you like exercising? | • *How old are you ? in which year at school you are? Do you have any brothers and sisters? What is your favourite subject?*<br><br>• *Which type of exercise is your favourite? Basketball, ballet, swimming?*<br><br>• *What is it that you like / dislike about exercising?* | • Make participant relax and feel comfortable with talking and opening up.<br>• Build rapport. |
| Transition Questions | • Talking about your back? Can you tell me how you have been diagnosed ?<br><br>• What was your thoughts about it ? | • *When did you first became aware of your back condition? How is that?*<br><br>• *How did you feel ?* | • Start to guide the interview towards experiences of patient with scoliosis<br><br>• To get an idea of their thoughts of having scoliosis.<br><br>• Explore feeling and perceptions of having scoliosis. |

| | | | |
|---|---|---|---|
| concept elicitation | • Thinking about your back, do you or have you ever had any difficulties?<br><br>• Do you ever take any painkillers or medicine for your back? | • *Are there any further issues that you can think of?*<br>• *(For example, your body is hurting you; pain at your joints; You feel pain during or after doing exercise/PE; chest pain; difficult breathing, decrease in the range of your movement? How is that?*<br>• *Can you think of other parts of your body that are affected because of your back? How do you feel it is affected? why?*<br>• *How many times do you take pain medications? Is it effective?*<br>• *What do you do to ease pain other than taking medications?*<br>• *If we want to measure the pain that you usually have. How much would you give it, in a scale from 0 to 10 ?* | • To explore effects of scoliosis on participant's body functions.<br>• To explore the effect of scoliosis on body structures. |
| | • Can you tell a bit more about any difficulties you have with your back? Which aspects of your daily life does it affects?<br><br>• Can you explain for me if and how your back affects you at school and activities linked to school? | • *Walking to school/catching the bus, climbing stairs, carrying bag.*<br>• *Sitting, standing, bending, kneeling.*<br>• *Studying/doing your homework, working in lab/art/music class.*<br>• *Do you participate in PE/school sports? How do you feel about doing exercise or sports with your classmates?*<br>• *Have you ever had days off school due to your back issue? Why?* | • To explore the effect of scoliosis on activities and participation (daily life activities).<br>o Mobility<br>o General task demand<br>o Major life areas |
| | • Does your back affect your ability to do activities with family or friends? | • *How is it affect your visiting relatives ? or friends?*<br>• *Does it affect your ability to go out with family? or friends?*<br>• *Does your back affect your ability to do things around home? For example : playing sport, Joining a game ?* | • To explore the effect of scoliosis on activities and participation (social life).<br>o Interpersonal interactions and relationships<br>o Community, social and civic life |

| | | | |
|---|---|---|---|
| | • Does your curve/back affect your ability to do things at home at all? | • *How your back affect you when taking care of yourself e.g., eating, bathing, toileting, dressing?*<br>• *Tidying up your room, helping family in preparing meals.*<br>• *Assisting others*<br>• *Taking care of pets/plants.* | • To explore effect of scoliosis on activities and participation<br>o Self-care.<br>o Domestic life |
| | • Can you describe your feelings when you have an issue with your back, how does this make you feel? | • *Does it worry you at all?*<br>• *Do you know what it is specifically that worries you? ( Fear of pain, difficulty preforming your homework, your scores in school less than usual, limited in activities that you can do).*<br>• *Does this affect your energy, (you feel tired)?*<br>• *Does your back condition ever make you feel down or discouraged, afraid, angry, or anxious ?*<br>• *Does is it affect your ability to sleep?* | • To explore the effect of scoliosis on mental function<br>o Emotional function<br>o Energy level<br>o Sleep function<br>o Personality function |
| | • How do you feel about the shape of your back? | • *Do you think your back shape is affecting your life? How is that ?*<br>• *Do you think it is limiting your ability to do things? How ?*<br>• *Do you think about your back when getting ready for sports / changing into uniform or sports kit at school / away from home or with others?*<br>• *Do you think it is affecting your relationship with other?* | • To explore the effect of scoliosis on mental function (body image)?<br>• Explore participants perception on their back shape. |
| | • Can you tell me what management/treatment that you have received for your back?<br><br>Can you describe the help that you received during treatment/management? | • *What was the treatment you received for your back?*<br>• *How do you think/feel about it ?*<br>• *Do you have an idea about other options of treatments you might receive for your back ? what is it ?*<br>• *How do you feel if you have the same treatment again?*<br><br>• *Which type of help/support have you received ? from whom? (Support from family, friends, peers, health professionals ( doctors,* | • To explore the experience of participants with their management and their satisfaction.<br><br>• To explore the effect of environmental factors on scoliosis |

| | | | |
|---|---|---|---|
| | | *nurses, physiotherapists, other healthcare providers?*<br>• *What do you think about it?*<br>• *Have you ever seen a physiotherapist or other therapist (e.g., osteopath/chiropractor)? What did you do with them? Did it help?*<br>• *Did you feel able to continue being active after seeing the physiotherapist?*<br>• *Is there anything you feel would improve your experience of living with scoliosis? what is it?* | |
| | • Now, we have reached the end of first part of this interview | • *Do you need a break?* | |
| Cognitive debriefing | • For the second part of our discussion, I want to see if the SRS-22r questionnaire is suitable for young people like you<br>• I will read the question one by one to you. Then please tell me, what do you understand the question is asking? | • *Have you completed this questionnaire before?* | • To test the relevance of SRS-22r items to individuals with AIS<br>• To assess if the SRS-22r items are understandable by individuals with AIS.<br>• To assess if the is SRS-22r is comprehensive. |
| | 1. Which one of the following best describes the amount of pain you have experienced during the past 6 months? -Response options are: None, Mild, Moderate, Moderate to severe, Severe. | • *Can you tell me in your words, what this question is asking you about?*<br>• *Do you think this question is Important to you ? why?*<br>• *Is there any word that is not clear to you ? what is it?*<br>• *Can you understand the answers? Is there any word that is not clear to you? what is it?* | |
| | 2. Which one of the following best describes the amount of pain you have experienced over the last month? -Response options are: None, Mild, Moderate, Moderate to severe, Severe | • *Can you tell me in your words, what this question is asking you about?*<br>• *Do you think this question is Important to you ? why?*<br>• *Is there any word that is not clear to you ? what is it?*<br>• *Can you understand the answers? Easy to choose the answer?* | |

| | | |
|---|---|---|
| | 3. During the past 6 months have you been a very nervous person? - Response options are: None of the time, A little of the time, Some of the time, Most of the time, All of the time. | • *Do you think this question is relevant to you? or Important to you ? why?*<br>• *Is there any word that is not clear to you ? what is it?*<br>• *Was it easy or difficult to choose that particular answer? Why?* |
| | 4. If you had to spend the rest of your life with your back shape as it is right now, how would you feel about it? - Response options are : Very happy, Somewhat happy, Neither happy nor unhappy, Somewhat unhappy, Very unhappy | • *What do you think the question is asking you about?*<br>• *Is it Important to you ?why?*<br>• *Was it easy or difficult to choose your answer? Why?*<br>• *Do you find this question too personal/ or embarrassing? Why?* |
| | 5. What is your current level of activity? Response options are : Bedridden, Primarily no activity, Light labour and light sports, Moderate labour and moderate sports, Full activities without restriction. | • *Do you understand the question? is it important to you?*<br>• *Are there any words that are not clear to you? What is your thought about these words "labour", "Bedridden", "restrictions"?*<br>• *Was it easy or difficult to choose your answer? Why?* |
| | 6. How do you look in clothes? - Response options are : Very good, Good, Fair, Bad, Very bad | • *Do you understand the question? is it important to you?*<br>• *Was it easy or difficult to choose your answer? Why?* |
| | 7. In the past 6 months have you felt so down in the dumps that nothing could cheer you up? -Response options are :Very often, Often, Sometimes Rarely, Never | • *Do you think this question is relevant to you?*<br>• *Are there any words that are not clear to you? Do you understand word "dump "? What does it mean?*<br>• *Do you find this question too personal/ or embarrassing? Why?*<br>• *Was it easy or difficult to choose your answer? Why?* |
| | 8. Do you experience back pain when at rest? - Response options are : Very often, Often, Sometimes, Rarely, Never | • *Do you think this question is important to you?*<br>• *Was it easy or difficult to choose your answer? Why?* |

| | | |
|---|---|---|
| | 9. What is your current level of work/school activity? -Response options are : 100% normal, 75%normal 50% normal, 25% normal, 0% normal | • *What went on in your mind when you were asked the question?*<br>• *is there any word that you don't understand?*<br>• *What is your answer? Would you explain to me how you come out with that answer ?* |
| | 10. Which of the following best describes the appearance of your trunk; defined as the human body except for the head and extremities?<br>-Response options are : Very good, Good, Fair, Poor, Very Poor | • *What do you think the question is asking you about?*<br>• *Is there any word that you don't understand?*<br>• *Was it easy or difficult to choose your answer? Why?* |
| | 11. Which one of the following best describes your pain medication use for back pain?<br>-Response options are : None, Non-narcotics, weekly or less (e.g., aspirin, Tylenol, Ibuprofen) , Non-narcotics daily, Narcotics weekly or less (e.g. Tylenol III, Lorcet, Percocet), Narcotics daily. | • *What went on in your mind when you read the question?*<br>• *Is there any word that is not clear to you ? what is it?*<br>• *What does the word "* Non-narcotics" *means to you?*<br>• *What is your answer? Would you explain to me how you came out with that answer ?* |
| | 12. Does your back limit your ability to do things around the house? -Response options are: Never, Rarely, Sometimes Often, Very Often | • *What do you think about this question? Is it important to you ?*<br>• *What about the answers? Was it easy or difficult to choose your answer? Why?* |
| | 13. Have you felt calm and peaceful during the past 6 months?<br>-Response options are: All of the time, Most of the time, Some of the time, A little of the time, None of the time. | • *What do you think the question is asking you about?*<br>• *Is there any word that is not clear to you ? what is it?*<br>• *Was it easy or difficult to choose your answer? Why?* |
| | 14. Do you feel that your back condition affects your personal relationships? -Response options are: None, Slightly, Mildly, Moderately, Severely | • *Do you think this question is important to you?*<br>• *Is there any word that you do not understand?*<br>• *Was it easy or difficult to choose your answer? Why?* |
| | 15. Are you and/or your family experiencing financial difficulties because of your back? | • *What went on in your mind when you read the question?* |

| | | |
|---|---|---|
| | -Response options are: Severely, Moderately, Mildly Slightly, None. | • *Do you find this question too personal/ or embarrassing? Why?*<br>• *Was it easy or difficult to choose your answer? Why?* |
| | 16. In the past 6 months have you felt down hearted and blue? -Response options are: Never Rarely, Sometimes, Often, Very often. | • *What do you think the question is asking you about?*<br>• *Is there any word that you do not understand? what about "down hearted', "blue"?*<br>• *Do you find this question too personal/ or embarrassing? Why?*<br>• *Was it easy or difficult to choose your answer? Why?* |
| | 17. In the last 3 months have you taken any days off of work, including household work, or school because of back pain? -Response options are : 0 days 1 day,2 days,3 days, 4 or more days. | • *Is this question important/relevant to you?*<br>• *What is your answer? Was it easy or difficult to choose your answer?* |
| | 18. Does your back condition limit your going out with friends/family? -Response options are : Never, Rarely, Sometimes, Often, Very often | • *Is this question important/relevant to you?*<br>• *What is your answer? Was it easy or difficult to choose your answer?* |
| | 19. Do you feel attractive with your current back condition? -Response options are :<br>Yes, very<br>Yes, somewhat, Neither attractive nor unattractive<br>No, not very much<br>No, not at all | • *Is this question important/relevant to you?*<br>• *Is there any word that you do not understand?*<br>• *Do you find this question too personal/ or embarrassing? Why?*<br>• *What is your answer? Was it easy or difficult to choose your answer?* |
| | 20. Have you been a happy person during the past 6 months? -Response options are :<br>None of the time, A little of the time, Some of the time, Most of the time, All of the time | • *Is this question important/relevant to you?*<br>• *Do you find this question too personal/ or embarrassing? Why?*<br>• *What is your answer? Was it easy or difficult to choose your answer?* |
| | 21. Are you satisfied with the results of your back management? | • *What do you think the question is asking you about?* |

| | | | |
|---|---|---|---|
| | -Response options are :<br>Very satisfied, Satisfied<br>Neither satisfied nor unsatisfied, Unsatisfied, Very unsatisfied | • *Is this question important/relevant to you?*<br>• *Are there any words that you don't understand?*<br>• *What is your answer? Was it easy or difficult to choose your answer?* | |
| | 22. Would you have the same management again if you had the same condition?<br>-Response options are :<br>Definitely yes, Probably yes, Not sure, Probably not, Definitely not | • *Do you think this question is important to you?*<br>• *What is your answer? Was it easy or difficult to choose your answer?* | |
| | • Do you think the questionnaire covers all areas which are important to you ? | • *Can you think of other areas that should be included in the questionnaire?* | • To assess if there are any additional areas need to be covered. |
| Conclusion | • That's all the questions, is there anything else you would like to tell me about your back or the questionnaire.<br>• | •<br>• *Is there anything you would like to ask regarding the analysis of the data or the next steps of the process?* | • Ensure the participant is comfortable with what has been discussed. |
| | • Thank you for participating in this study, I really appreciate your time and input. | | |
